# Evaluating inequalities in out-of-pocket expenditures across inpatient and outpatient services in Honduras

**DOI:** 10.1101/2025.03.24.25324567

**Authors:** Carlos Rojas-Roque, Rodrigo Vargas-Fernández, Akram Hernández-Vásquez

**Affiliations:** Centre for Health Economics, University of York, Heslington, York, YO10 5DD, UK; Epidemiology and Health Economics Research (EHER), Universidad Científica del Sur, Lima, Peru; Centro de Excelencia en Investigaciones Económicas y Sociales en Salud, Vicerrectorado de Investigación, Universidad San Ignacio de Loyola, Lima, Peru

**Keywords:** Out-of-pocket expenditure, inequality, access to healthcare, Honduras

## Abstract

Out-of-pocket (OOP) healthcare expenditures are a major barrier to achieving universal health coverage in low- and middle-income countries (LMICs). In Honduras, OOP payments constitute a significant burden, particularly for vulnerable populations. However, evidence on the socioeconomic distribution of these expenditures and their determinants remains scarce. This study aims to examine the socioeconomic inequalities in OOP healthcare expenditures in Honduras, focusing on outpatient and inpatient services. It seeks to identify key sociodemographic factors contributing to these disparities to inform equitable health care financing policies. Data from the 2019 ENDESA/MICS survey was used, covering 10,998 individuals for outpatient and 3,277 for inpatient services. Concentration curves (CC) and the Wagstaff concentration index (CI) were used to measure inequality in OOP expenditures. An econometric decomposition analysis of the CI was performed to identify the contribution of sociodemographic factors. The findings indicate that OOP expenditures for both outpatient (CI = 0.213) and inpatient services (CI = 0.218) are disproportionately concentrated among wealthier individuals. Education and place of residence were primary drivers of inequality, with rural residents and those without insurance experiencing greater financial burdens. The study highlights significant socioeconomic inequalities in OOP healthcare expenditures in Honduras. Policy interventions targeting financial protection for lower-income and rural populations are crucial to advancing equitable healthcare access.

## Introduction

Out-of-pocket (OOP) healthcare expenditures represent a significant barrier to achieving universal health coverage (UHC) [1]. OOP payments, defined as direct payments made by individuals at the point of health service, report high levels in low- and middle-income countries (LMICs). For instance, in 2018, OOP health spending accounted for nearly 40% of total health expenditures in LMICs [2]. The reliance on OOP often leads to financial hardship, particularly for vulnerable populations, and can exacerbate socioeconomic inequalities, disproportionately affecting lower-income households lacking adequate financial protection [3].

In Honduras, the healthcare system is characterised by a dual structure of public and private providers. The public sector is composed by the Ministry of Health, which provides services to 60% of the population, and by the Honduran Institute of Social Security (IHS) which secures services to 12% of the population [4]. The private sector is composed of profit and non-profit organizations, which provides services to 10% of the population [4]. The remaining 18% of the population is estimated to not have access to health services [4]. This lack of effective coverage is reflected in the high levels of OOP expenditures, which account for 48.7% of total health expenditure—significantly higher than the Latin American and Caribbean average of 28.2% [5].

The burden of OOP expenditures is further illustrated by data from the 2019 National Demographic and Health Survey/Multiple Indicator Cluster Survey (ENDESA/MICS 2019). In 82% of cases, the costs of outpatient services incurred over the past 30 days were borne directly by the patients themselves [6]. The average percapita OOP expenditure for these services was 743 lempiras (approximately 30 USD in 2019) [6,7]. In contrast, over the past 12 months, 17.7% of inpatient services were provided by private institutions [6], with an average per-capita OOP expenditure of 6,971 lempiras (approximately 282 USD in 2019) [6,7]. Such high OOP costs exacerbate existing disparities in healthcare access and outcomes, posing a significant concern for healthcare financing and equity in Honduras.

Understanding the socioeconomic distribution of OOP healthcare expenditures and the factors contributing to these disparities is crucial for informing policies aimed at enhancing healthcare access and promoting financial equity. However, there is currently limited evidence describing the extent and nature of these inequalities in OOP expenditures in Honduras. This paper seeks to address this gap by employing concentration curves (CC) and the Wagstaff concentration index (CI) to systematically examine disparities in OOP expenditures across both inpatient and outpatient services in Honduras. Given that OOP spending can vary significantly between different types of health services [8,9], the inclusion of both inpatient and outpatient OOP expenditures is anticipated to yield comprehensive insights with significant policy implications for alleviating the financial burden on households. Furthermore, a decomposition analysis of the CI is conducted to identify the key sociodemographic determinants contributing to these inequalities. The findings of this study are expected to offer valuable intuition into the financial challenges faced by diverse population segments and to inform ongoing policy discussions on equitable healthcare financing in Honduras.

## Materials and methods

### Study setting

Honduras is a lower-middle-income country (LMIC) within the Central American region. As of 2023, the country has a population of 10.6 million and a Gross Domestic Product (GDP) per capita of approximately US$3,232 [2]. Honduras struggles with high poverty rates: in 2023, 51.9 percent of the population was poor, based on a poverty threshold of US$6.85 per capita per day (2017 purchasing power parity) [10]. Health indicators reflect these socioeconomic challenges, with a life expectancy at birth of 71 years (2022) [2], an infant mortality rate of 13.8 per 1,000 live births [2], and a maternal mortality ratio of 72 deaths per 100,000 live births [2]. The burden of disease is dominated by interpersonal violence, neonatal disorders, and ischemic heart disease, which are the leading causes of death and disability [5]. Notably, communicable diseases such as tuberculosis, dengue, and Zika remain persistent public health concerns [5], underscoring the complex epidemiological landscape of the country.

### Data source

This study used the 2019 cross-sectional ENDESA/MICS, carried out by the National Institute of Statistics and the Health Secretariat in Honduras. The survey collects nationwide representative information on fertility, general and reproductive health, housing indicators, nutritional status of children, infant mortality, HIV/AIDS, domestic violence, morbidity, use of services, health expenses and demographic indicators of the Honduran population. The ENDESA/MICS 2019 sample design is probabilistic, stratified and two-staged. Further methodological details can be freely accessed in the survey website [6].

### Sample size

The eligible population was defined as individuals aged 4 years or older who were residents of urban or rural locations. For the outpatient services, we included only those individuals who paid a health professional to treat their health problem during the last 30 days. For the inpatient services, we included only those individuals who reported expenditure for the hospitalisation during the past 12 months. The final sample size is composed of 10,998 individuals for the outpatient services and 3,277 individuals for the inpatient services. The flowchart of the sample size of the study is reported in Fig 1.

**Figure 1.**
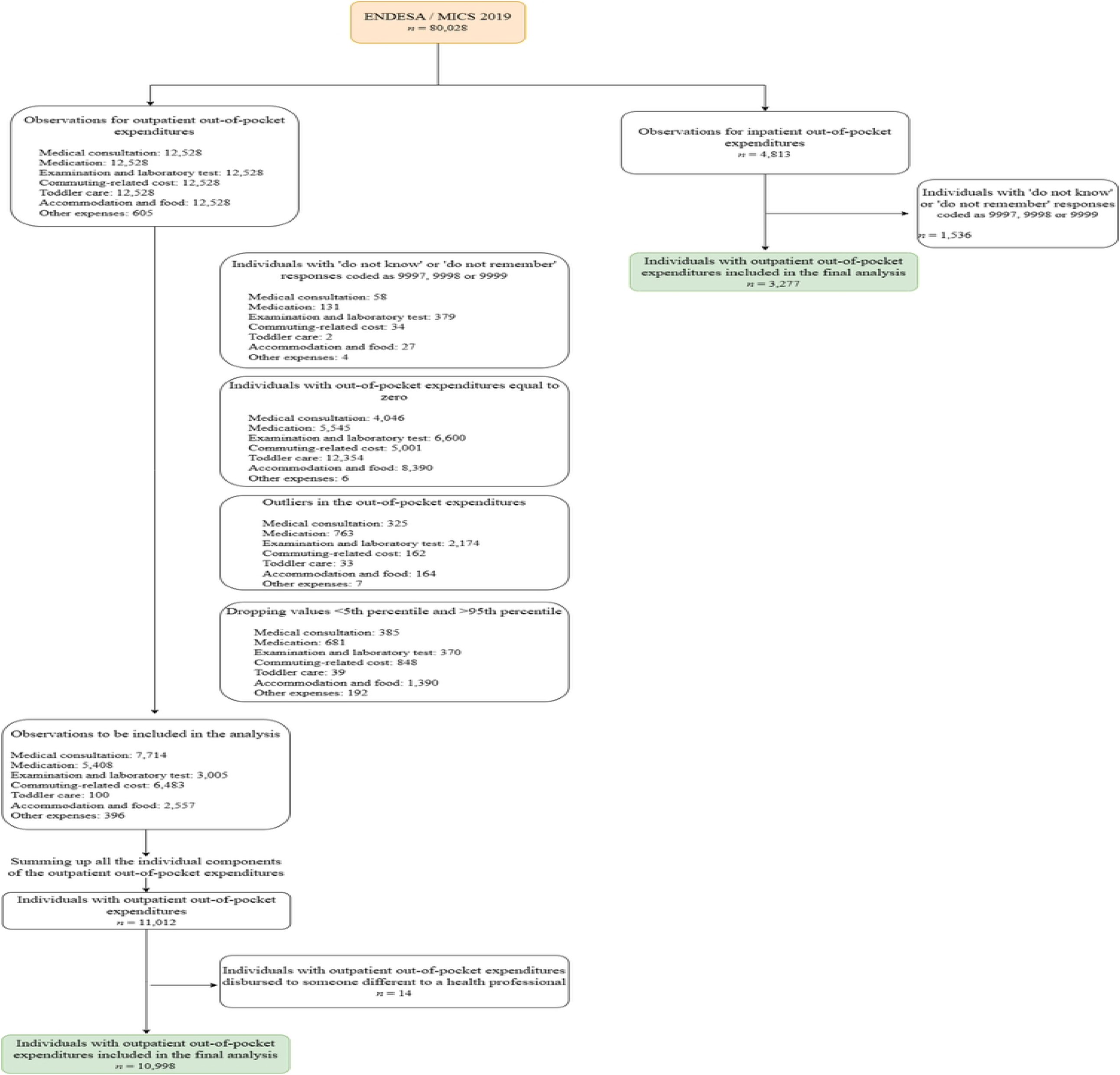
Flowchart of the study participants.

### Outcome variables

The outcomes of the study are the outpatient and the inpatient OOP expenditure. Both outcome variables were measured in Lempiras, yet reported in US dollars as for 2019. We used the average exchange rate between 10 June 2019 and 14 December 2019 (coinciding with the ENDESA/MICS 2019 fieldwork period) published by the Central Bank of Honduras [7].

To calculate the outcome variables, we used the ‘Household questionnaire’. For the outpatient OOP, the variable was constructed by adding all the payments (the name of the variables in the dataset are displayed within square brackets []) disbursed by medications [AE18], medical consultation [AE17], examinations and laboratory tests [AE19], commuting-related cost [AE22A], toddler care [AE22B], accommodation and food [AE22C] and other expenses [AE24]. For the inpatient out-of-pocket expenditures, the variable was constructed by adding the payment disbursed by hospitalisation, including medical consultation, medications, examinations and laboratory tests [HE8], commuting-related cost [HE11A], toddler care [HE11B] and accommodation and food [HE11C]. The reference period for the outcomes is 30 days for outpatient out-of-pocket expenditures and 12 months for the inpatient out-of-pocket expenditures.

### Covariates

Based on previous literature,[11,12] we included the following covariates: sex (man / woman), age (0-14 years / 15-64 years / 65-74 years / 75 years or older), educational level (no education / basic / secondary / university), place of residence (urban / rural), health insurance coverage (yes / no), economic status (poorest quintile / second quintile / middle quintile / fourth quintile / richest quintile) and region (central / western / northern / Mosquitia / eastern / southern).

### Statistical analysis

All analyses were performed using STATA v.18.0, taking into account the survey’s sampling design and expansion factors. Socio-demographic variables were described by using weighted proportions.

To assess socioeconomic inequality in OOP expenditures, we estimated the concentration curves (CC) and the Wagstaff concentration index (CI). CC plots the cumulative percentage of the health variable (*y-*axis) against the cumulative percentage of the population ranked by socioeconomic status, starting with the poorest and ending with the richest (*x-*axis). Hence, if all individuals, regardless of their socioeconomic status, possess identical levels of health, the concentration curve will resemble a diagonal line starting from the lower left and ending at the upper right, referred to as the line of equality. Conversely, if health indicators skew towards higher (lower) values among those with lower income, the concentration curve will deviate above (below) the line of equality. The greater the distance between the curve and the line of equality, the more pronounced the concentration of health outcomes among individuals with lower or upper socio-economic status [13,14].

On the other hand, CI is defined as twice the area between the line of equality and the CC. Thus, in the case of total equality, the CI takes the value of zero. The CI takes a negative value (positive value) when the CC lies above (below) the line of equality, indicating disproportionate concentration of the health among the poorest (richest) ones [14]. Based on Koolman and van Doorslaer [15], by multiplying the CI by 75, one can ascertain the proportion of the health variable that would require redistribution from the wealthier half to the less affluent half of the population, assuming health inequality privileges the affluent, in order to achieve a distribution yielding an index value of zero.

For computational convenience, the CI can be defined in terms of the covariance between the healthy variable and the fractional rank in the living standard distribution:

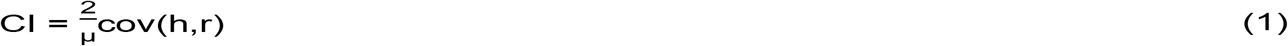

In Equation (1), h is the health variable, r is the rank of the living standard variable and μ is the mean of the health variable.

Lastly, we decompose the CI to estimate the contribution of each covariate to the inequality in the out-of-pocket expenditures. We can express the model of out-of-pocket expenditures (y) as follows:

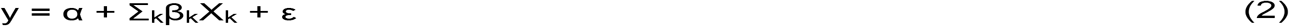

In Equation (2), α represent the intercept, X represent the covariates and β represent its regression coefficient, ε represent the stochastic term of error. Equation (2) was estimated using ordinary least squares (OLS). Using the regression equation, CI can be decomposed as follows:

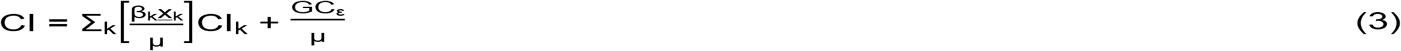

In Equation (3), μ represent the mean of y (inpatient and outpatient out-of-pocket expenditures), <u>x</u>_k_ represent the mean of X_k_, CI_k_ is the concentration index for X_k_ while GC_ε_ is the generalised concentration index for the error term, ε. Based on Equation (3), CI is equal to a weighted sum of the concentration indices of the k covariates, where the weight for X_k_ is the elasticity of y with respect to 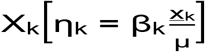.

The residual component, as indicated by the final term, signifies the income-related disparity in health that remains unaccounted for by systematic variations in the regressors based on income. This residual should tend toward zero in a fully properly specified model [14].

The decomposition analysis reports elasticity, concentration, contribution, and percentage of contribution for each covariate. Elasticity indicates the change in the occurrence of the outcome of interest (out-of-pocket expenditure) corresponding to a one-unit alteration in the independent variable. A positive or negative elasticity signifies an increased or decreased amount of out-of-pocket expenditure with a change in the covariate, respectively [14]. The concentration index reflects the degree of concentration of variables concerning quintiles of household income per capita. A positive or negative value indicates a higher out-of-pocket expenditure among the wealthiest or poorest households, respectively [14]. Finally, contribution and percentage contribution denote the absolute and relative contributions of each covariate to the overall socioeconomic inequality in the out-of-pocket expenditure. A positive or negative contribution or percentage contribution for a covariate leads to an increase or decrease in observed socioeconomic inequality, respectively [14].

### Ethic statement

This study did not require the approval of an institutional ethics committee since all the databases are fully anonymized and are freely and publicly available from the following website: https://mics.unicef.org/news/just-released-honduras-2019-survey-findings-and-datasets.

## Results

Table 1 describes the sociodemographic characteristics of the population (*n*=10,998). Most of the population for both outpatient and inpatient out-of-pocket expenditures are women (59.2% and 63.1% respectively). Nearly half of the outpatient services out-of-pocket expenditures sample are between 15 and 64 years old, while two thirds of the inpatient services out-of-pocket expenditures sample are reported by individuals aged between 15 and 64 years old. For both healthcare services, the majority of the individuals reported basic education (64% and 64.9%) and reported being from the rural area (56% and 54.4%). For both inpatient and outpatient services, the vast majority of the population reported not having health insurance coverage (85.1% and 88.8% respectively).

**Table 1.**
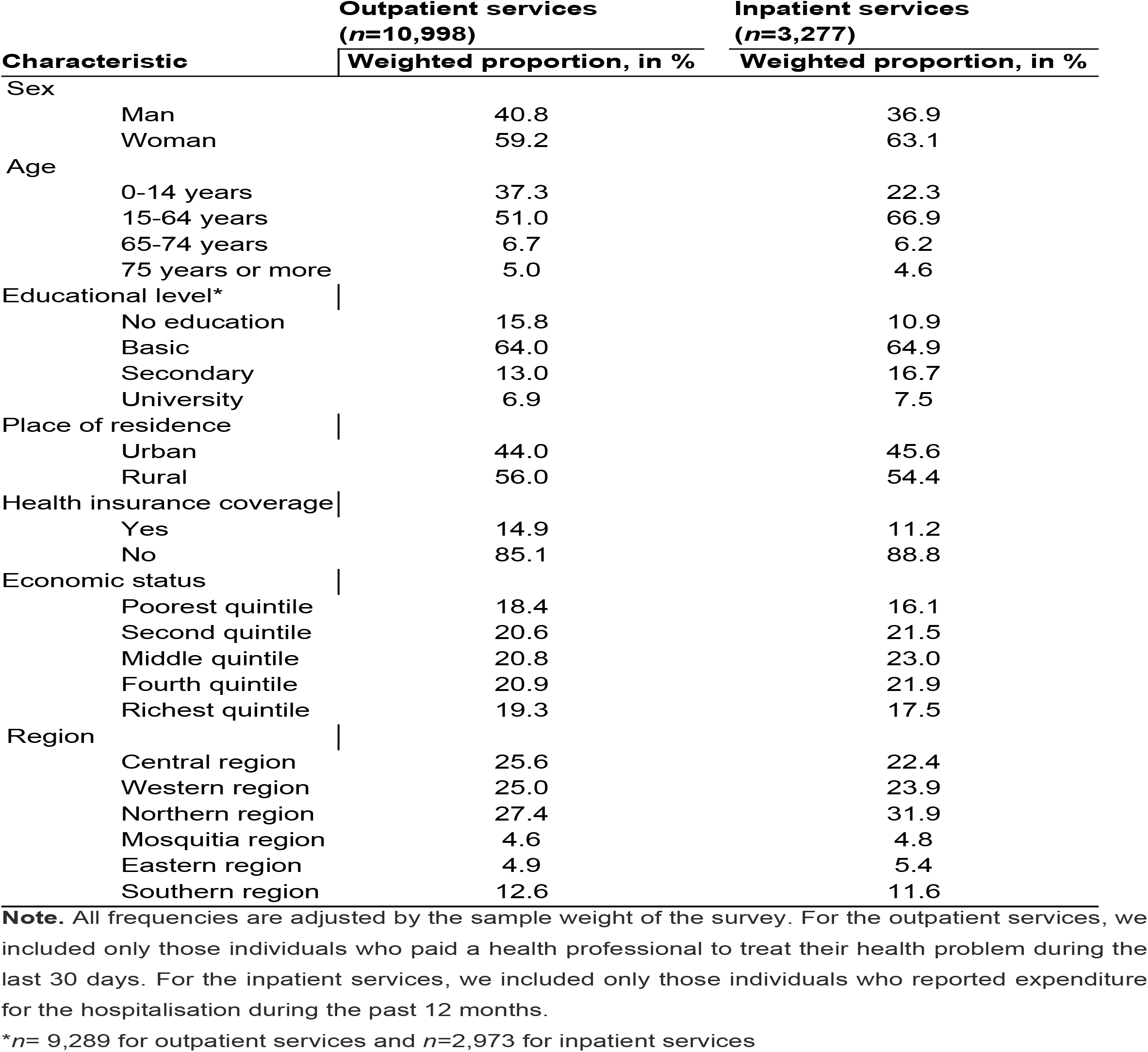
Characteristics of the population.

The Wagstaff CI for outpatient and inpatient services are reported in Table 2. Overall, the CI for outpatient services is 0.213. The positive sign of the CI reveals that the out-of-pocket expenditures in outpatient services are disproportionately concentrated among the wealthiest individuals. Following the approach proposed by Koolman and van Doorslaer [15], in order to achieve an equal distribution of the out-of-pocket expenditures, the proportion of out-of-pocket expenditures that would require redistribution from the wealthier half to the less affluent half of the population is 15.9. In other words, it is required that 15.9% of the wealthier half individuals redistribute their out-of-pocket expenditures to the less affluent half individuals to achieve a CI equal to zero.

**Table 2.**
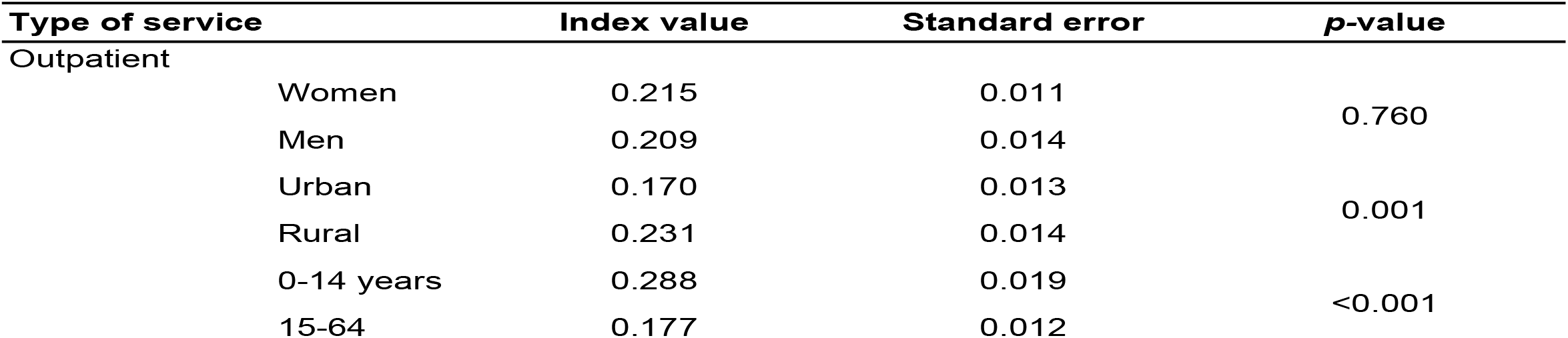
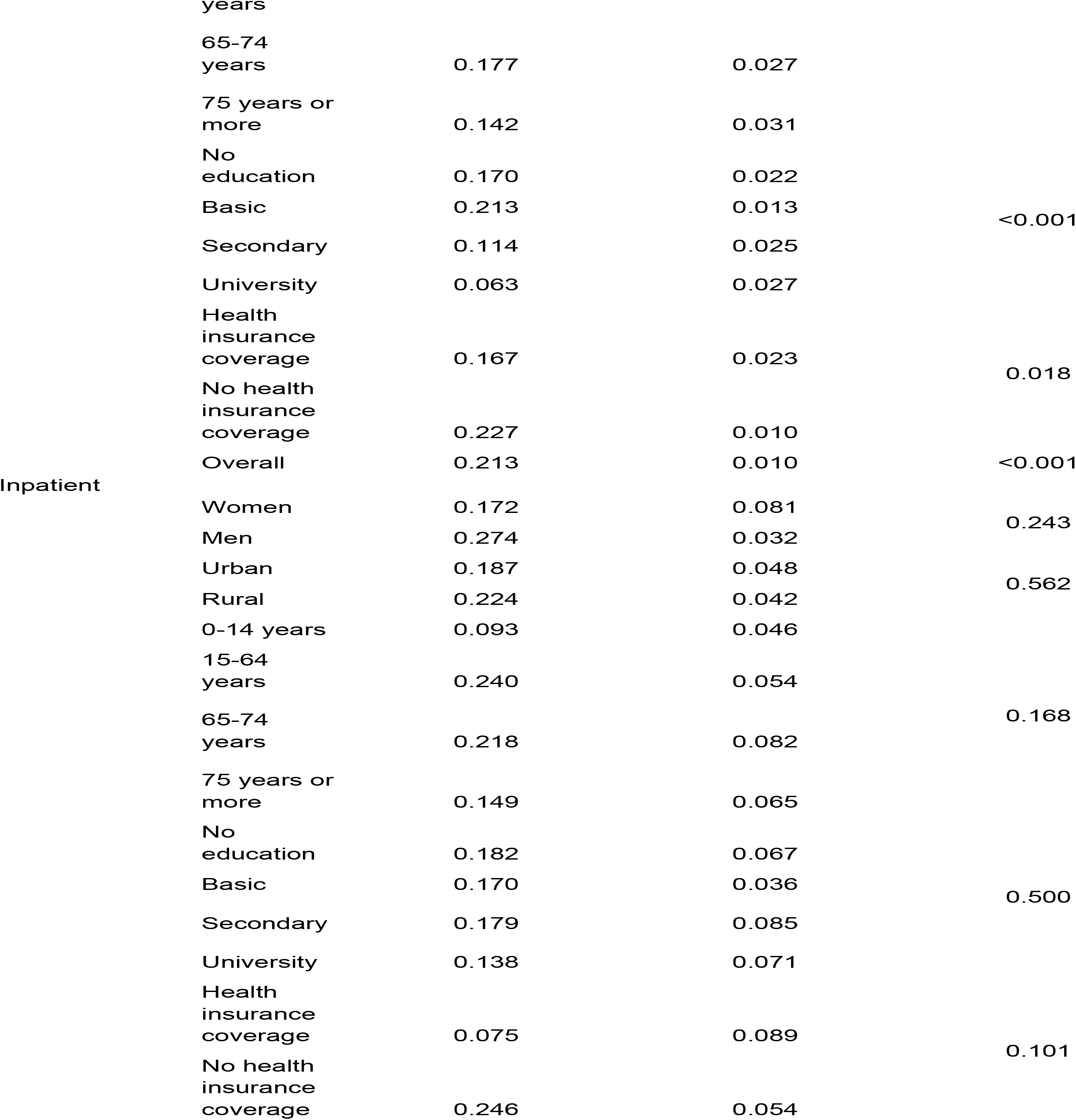
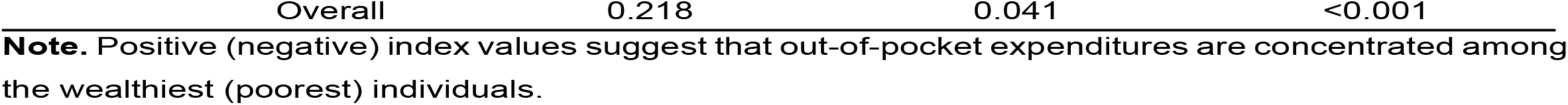
Wagstaff concentration index for outpatient and inpatient services in the out-of-pocket expenditures in Honduras.

| Type of service | Index value | Standard error | p-value |
| --- | --- | --- | --- |
| Outpatient |  |  |  |
| Women | 0.215 | 0.011 | 0.760 |
| Men | 0.209 | 0.014 |  |
| Urban | 0.170 | 0.013 | 0.001 |
| Rural | 0.231 | 0.014 |  |
| 0-14 years | 0.288 | 0.019 | <0.001 |
| 15-64 | 0.177 | 0.012 |  |

|  |  |  |  |  |
| --- | --- | --- | --- | --- |
|  | years |  |  |  |
|  | 65-74 years | 0.177 | 0.027 |  |
|  | 75 years or more | 0.142 | 0.031 |  |
|  | No education | 0.170 | 0.022 |  |
|  | Basic | 0.213 | 0.013 | <0.001 |
|  | Secondary | 0.114 | 0.025 |  |
|  | University | 0.063 | 0.027 |  |
|  | Health insurance coverage | 0.167 | 0.023 | 0.018 |
|  | No health insurance coverage | 0.227 | 0.010 |  |
|  | Overall | 0.213 | 0.010 | <0.001 |
| Inpatient | Women | 0.172 | 0.081 | 0.243 |
|  | Men | 0.274 | 0.032 |  |
|  | Urban | 0.187 | 0.048 | 0.562 |
|  | Rural | 0.224 | 0.042 |  |
|  | 0-14 years | 0.093 | 0.046 |  |
|  | 15-64 years | 0.240 | 0.054 |  |
|  | 65-74 years | 0.218 | 0.082 | 0.168 |
|  | 75 years or more | 0.149 | 0.065 |  |
|  | No education | 0.182 | 0.067 |  |
|  | Basic | 0.170 | 0.036 | 0.500 |
|  | Secondary | 0.179 | 0.085 |  |
|  | University | 0.138 | 0.071 |  |
|  | Health insurance coverage | 0.075 | 0.089 | 0.101 |
|  | No health insurance coverage | 0.246 | 0.054 |  |
| Overall | 0.218 | 0.041 | <0.001 |

**Note.** Positive (negative) index values suggest that out-of-pocket expenditures are concentrated among the wealthiest (poorest) individuals.

We do not find statistical differences in the CI for outpatient services in the out-of-pocket expenditures according to sex (*p*=0.760). However, differences in the CI were reported according to the place of residence (*p*=0.001), age (*p*<0.001), educational level (*p*<0.001) and health insurance coverage (*p*=0.017).

For the inpatient services, the overall CI is 0.218. Again, the positive sign of the CI denotes that the out-of-pocket expenditures in the inpatient services are concentrated among the wealthiest individuals. In order to achieve an equal distribution in the out-of-pocket expenditures, 16.3% of the wealthier half individuals have to redistribute their out-of-pocket expenditures to the less affluent half individuals [15]. We do not report statistical differences in the CI in any of the subgroups included in the analysis.

The CCs for both services are reported in Fig 2. Both curves lie below the equality line, confirming that out-of-pocket expenditures for outpatient services and inpatient services are disproportionately concentrated among the wealthiest individuals. In the Electronic Supplementary Material 1 we reported subgroups CCs for both services. For outpatient services, the CC for rural individuals and the CC for individuals without health insurance coverage are more distant from the line of equity compared with the CC for urban individuals and compared with the CC for individuals with health insurance coverage, respectively. The findings suggest that the inequality is greater among individuals living in rural areas and among individuals without health insurance coverage. The results confirm the statistical differences reported in Table 2 for this subgroup population. For inpatient services, the CC for rural individuals and urban individuals overlapped across the cumulative population sorted by the wealth index. The results suggest that no major differences in the inequality are reported between groups. The result is confirmed in Table 2 where no statistical differences were reported. Similarly, despite that the CC for those individuals without health insurance coverage tends to be more distant from the line of equity when compared with the CC for those individuals with health insurance, we do not find any statistical differences (see Table 2).

**Figure 2.**
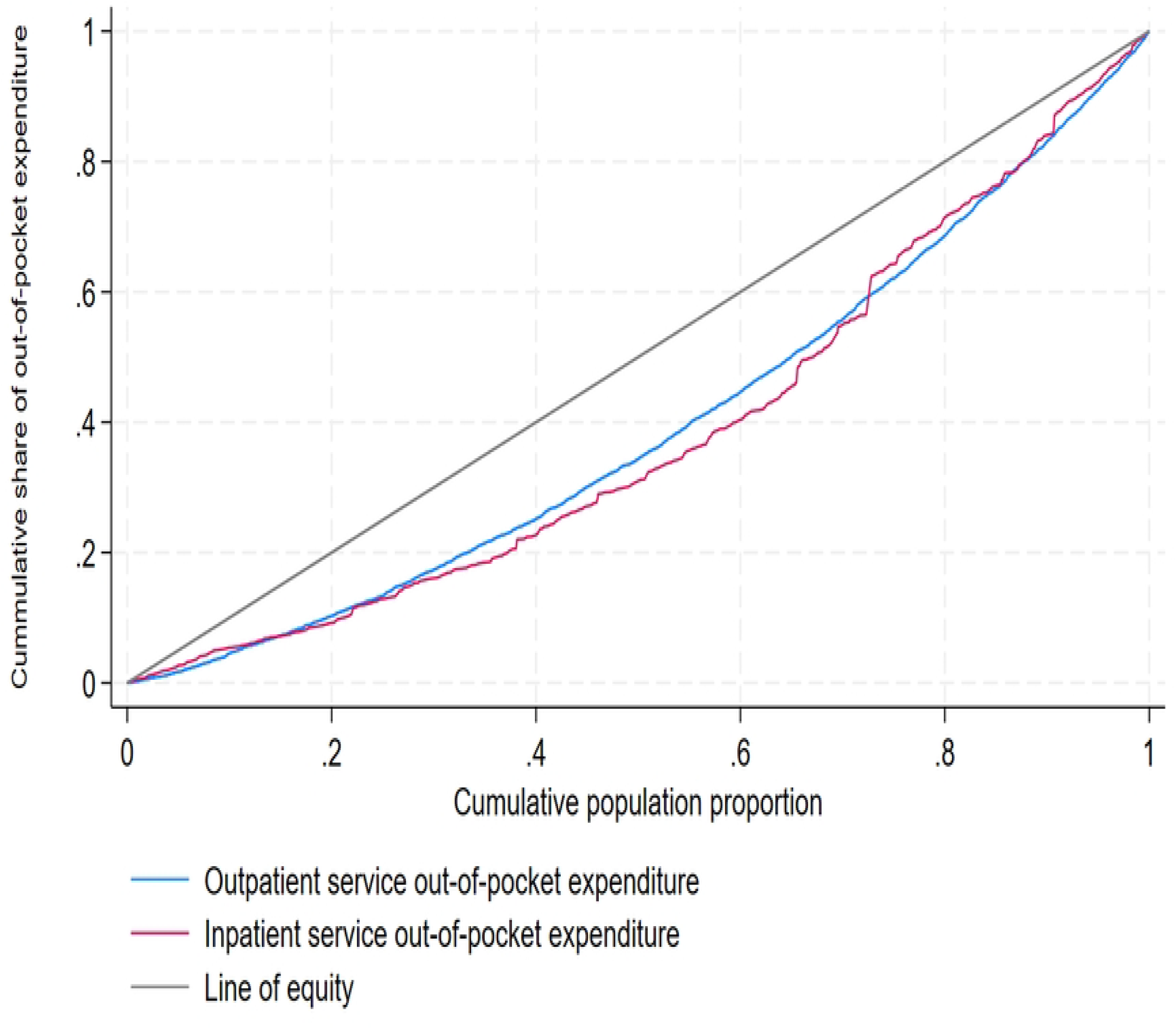
Concentration curves for outpatient and inpatient services out-of-pocket expenditures in Honduras.

Table 3 reports the decomposition of the Wagstaff CI for both services. For the outpatient out-of-pocket expenditures, the elasticity parameter reported a positive sign in all the covariates, except for place of residence and some regions from Honduras. The positive sign of the elasticity reflects an increased amount of out-of-pocket expenditure with a change in the covariate. For example, the elasticity for ‘no education’ is equal to 0.069, indicating that 1% of change of individuals from no education to basic education results in a 6.9% increase in out-of-pocket expenditure.

**Table 3.**
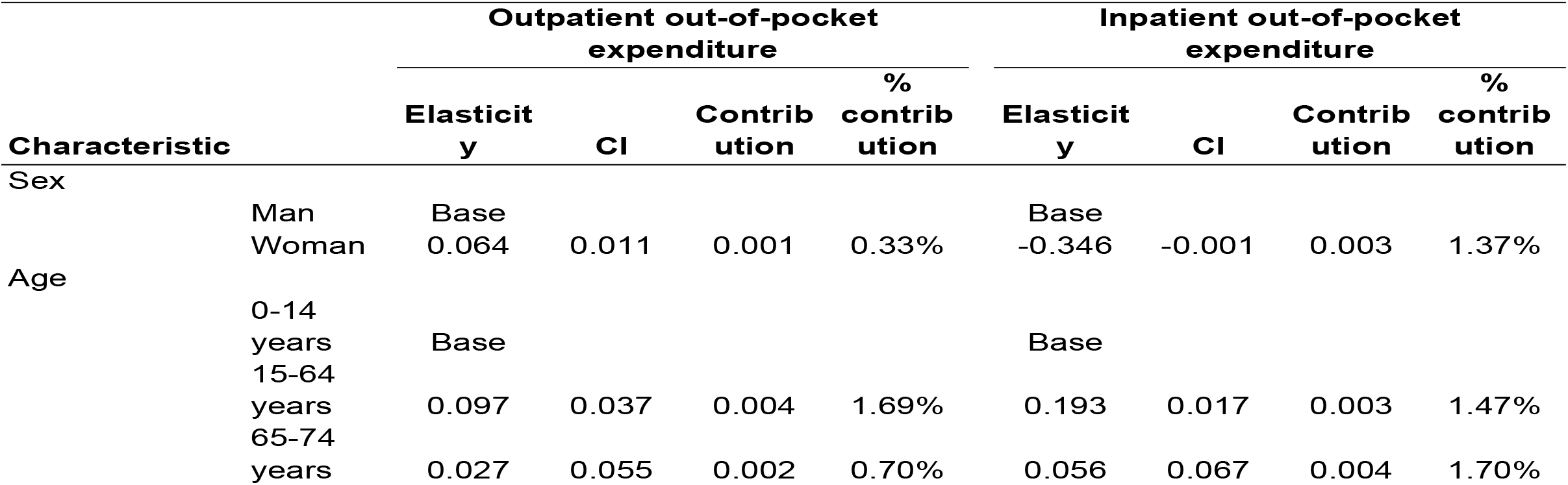
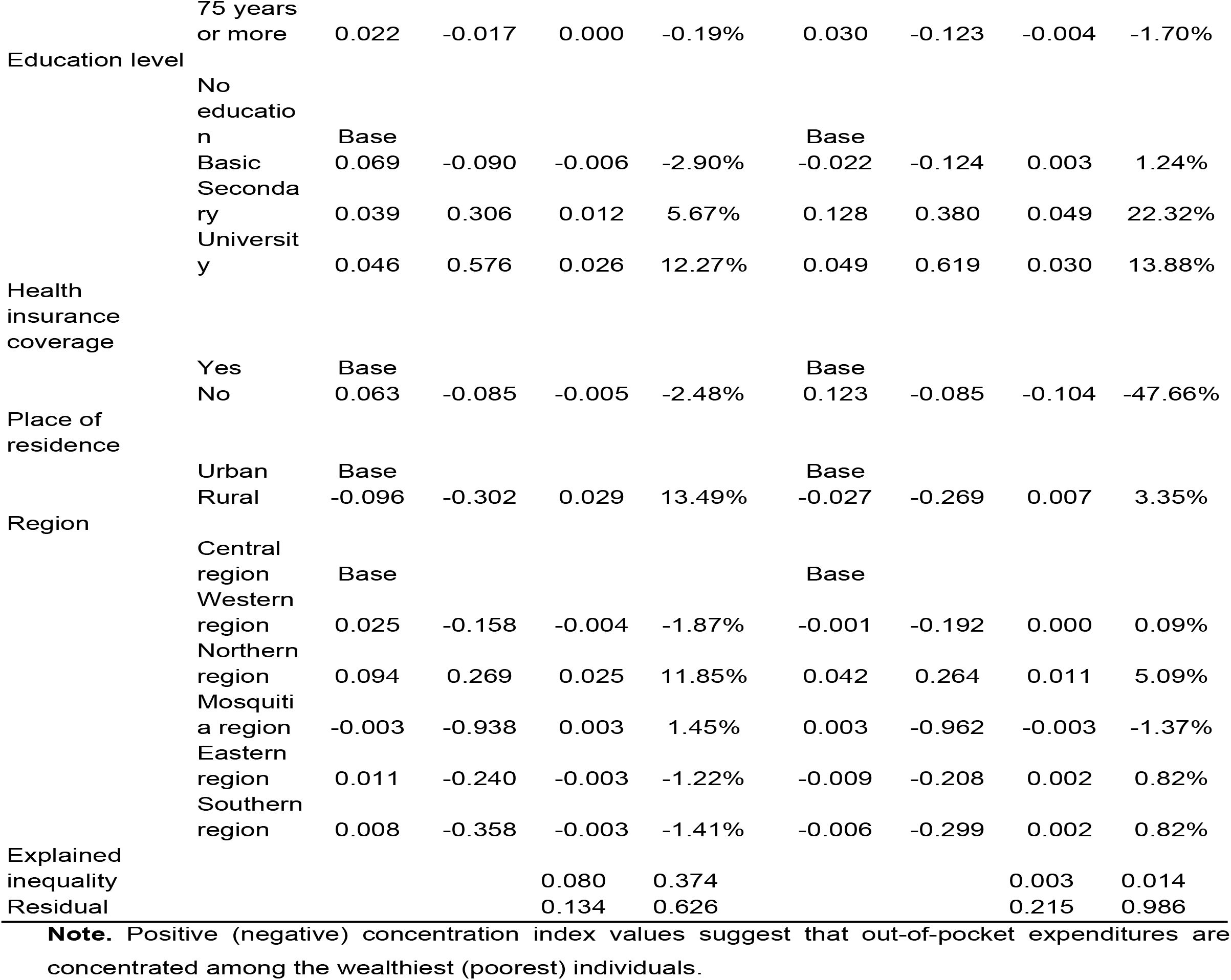
Decomposition of the Wagstaff concentration index for the outpatient and inpatient services out-of-pocket expenditures.

For the CI, there is a mix between positive and negative signs across the covariates. For example, the CI for being a woman is 0.001, suggesting that the out-of-pocket expenditures among women is concentrated among the wealthiest ones. On the other hand, the negative value of the CI for individuals living in the Southern region (-0.358) suggest that the out-of-pocket expenditures are concentrated among the poorest individuals. Lastly, for the contribution and percentage contribution of the covariates to the inequality in the out-of-pocket expenditure, we found that the main drivers of the inequality were the university education and urban place of residence.

For the inpatient out-of-pocket expenditures, the elasticity parameters reported a positive value in all the covariates, except for sex, basic education, place of residence and some geographic regions from Honduras. The positive elasticity suggests that changes in the covariates are associated with increased out-of-pocket expenditures. For instance, a 1% change of individuals from having health insurance to not having health insurance is associated with a 12.3% increase in out-of-pocket expenditures. For the CI, again there is a mix of signs across the covariates. As an example, the CI for being a woman is -0.001, suggesting that among women the out-of-pocket expenditures are concentrated among the poorest ones. However, women with university educational level reported a positive CI, which reflects that out-of-pocket expenditures are concentrated among the wealthiest women within this subgroup. Lastly, based on the contribution and percentage contribution, the main drivers of the inequality are educational level and place of residence.

## Discussion

This study aimed to evaluate inequalities in the out-of-pocket expenditures across inpatient and outpatient services in Honduras. Our results indicate that OOP expenditures for both outpatient and inpatient services are disproportionately concentrated among the wealthiest individuals, as evidenced by the positive values of the Wagstaff CI. The decomposition of the CI highlights the key contributors to inequality in OOP expenditures. For outpatient and inpatient services, education and place of residence emerged as the main drivers of disparity.

Our findings indicate that individuals with higher economic status bear a larger OOP healthcare costs, suggesting a tendency toward progressivity in the financing sustainability of Honduras’ health system. This observation is in line with previous research that found that OOP tends to be progressive and catastrophic expenditures tend to be concentrated among the rich when expenditures are assessed relative to consumption [16]. In fact, the ENDESA/MICS survey collects data on per capita OOP health expenses, effectively treating such expenditures as a consumption good. However, a limitation of this approach is its inability to determine whether the proportion of income spent on OOP health expenses is higher among poorer individuals (indicating regressive expenditures) or lower (indicating progressive expenditures). Addressing this limitation would require microdata that includes information on both OOP health expenditures and total household consumption or income—data that is not available in the ENDESA/MICS 2019 survey.

Two factors may reinforce the progressivity observed in the distribution of healthcare financing: the absence of a robust regulatory framework to oversee private providers and persistent barriers to accessing healthcare services. In Honduras the private healthcare subsector operates with autonomy and limited regulation [4]. This regulatory gap can lead to practices such as overcharging to offset inflationary pressures, over-provision of high-cost services (e.g., elective surgeries) at the expense of essential or preventive care—which are less profitable but critical for public health—and uneven distribution of services. Private providers tend to concentrate in urban or wealthier areas where profitability is higher, often neglecting rural or underserved regions. Consequently, higher OOP expenditures are disproportionately incurred by those who can afford to access these services.

Furthermore, structural and economic barriers exacerbate inequities in healthcare access. With poverty levels estimated at 51.9 percent of the population in 2023 [10] and inadequate infrastructure—such as limited road networks and transportation options—many individuals in remote areas face significant challenges in reaching healthcare facilities [17]. These barriers disproportionately deter low-income populations from seeking care, leaving only those with sufficient financial means to bear the associated OOP costs.

### Policy implications

The results underscore the urgent need for targeted policy interventions to address persistent inequalities in health care financing. In this context, the ongoing technical cooperation between the Pan American Health Organization (PAHO) and the Honduran Secretary of Health presents a significant opportunity to mitigate these disparities [18]. Key initiatives, such as the Strategy for Strengthening Integrated Health Services Networks (RISS), grounded in Primary Health Care (PHC) principles, alongside the Ministry of Health’s immediate and short-term action plan for 2024 and the National Health Re-Foundation Plan 2023-2030, aim to adopt a multisectoral approach to develop more holistic and inclusive health policies. These efforts prioritize addressing the needs of the most disadvantaged populations, ensuring that healthcare access and financial protection are extended to those who are often marginalized. However, the success of these initiatives will depend on robust monitoring and evaluation mechanisms. Implementing a comprehensive surveillance system is essential to track the evolution of critical health outcomes and assess trends in health inequalities over time. Such a system would enable policymakers to identify gaps, measure the impact of interventions, and adjust strategies to ensure sustained progress toward equity in healthcare financing and access.

Addressing OOP healthcare expenditures in Honduras requires a multisectoral approach, particularly in view of the key drivers of inequality—namely, education and place of residence. Collaborative efforts with the Ministry of Education could prove instrumental in this context. Research indicates that low health literacy is significantly correlated with higher OOP expenditures [19], primarily due to delayed healthcare-seeking behaviors and inadequate utilization of preventive services. Therefore, the implementation of nationwide health education campaigns aimed at enhancing health literacy may be of high relevance. Such campaigns should specifically target populations with lower levels of educational attainment. Moreover, effective execution of these initiatives would benefit from partnerships with the Ministry of Education. Integrating health literacy into school curricula could serve as a sustainable strategy to improve long-term outcomes. Additionally, deploying community health workers to deliver educational programs in rural areas could help bridge the gap in healthcare access. This comprehensive approach could potentially mitigate the financial burden of healthcare on vulnerable populations and reduce the observed disparities in OOP expenditures.

Furthermore, it is important to consider that certain health inequalities in Honduras may stem from the structure and organization of the national health system. The coexistence of two distinct sectors—public and private—necessitates the implementation of additional policies aimed at regulating costs within the private sector. In parallel, efforts to promote public-private partnerships could serve as a strategic measure to enhance equitable access to healthcare services. Such initiatives would not only contribute to reducing financial barriers but also help protect the population from excessive OOP expenditures.

### Strength and limitations

This study has several limitations that should be acknowledged. First, the cross-sectional design limits the ability to infer causality between OOP expenditures and the examined sociodemographic variables. Nonetheless, the findings indicate a strong correlation between OOP expenditures, education, and place of residence. Additionally, the reliance on self-reported data for OOP expenditures may introduce recall bias, particularly when respondents are asked to report the amounts spent on healthcare services. This potential bias could affect the accuracy of the results. Moreover, the decision to include only individuals who had incurred some level of OOP expenditure presents a limitation in terms of generalizability. When decomposing the CI, factors influencing OOP expenditures among those who did not participate in the study may have been overlooked, which may potentially provide a more comprehensive understanding of the underlying inequalities. Future research might consider employing qualitative methodologies to explore barriers to OOP expenditures more deeply. Such approaches could offer valuable insights for policymakers and researchers aiming to develop strategies to mitigate OOP inequalities effectively.

## Conclusion

Overall, our study highlights significant socioeconomic inequalities in OOP expenditures in Honduras, with wealthier individuals shouldering a greater financial burden. While outpatient services exhibit more pronounced inequality across sociodemographic subgroups, inpatient services appear to have a more uniform distribution. Addressing these disparities requires implementing comprehensive health policies focused on strengthening financial protection mechanisms and ensuring equitable access to healthcare, particularly targeting rural and lower socioeconomic status populations.

## Data Availability

The dataset used is publicy available from the following website: https://mics.unicef.org/news/just-released-honduras-2019-survey-findings-and-datasets

## Acknowledgements

The authors would like to thank anonymous revisions on previous versions of the manuscript

